# Traditional treatment of HIV and the role of family members as barriers to access to HIV care service or antiretroviral therapy among people living with HIV

**DOI:** 10.1101/2022.02.28.22270871

**Authors:** Nelsensius Klau Fauk, Lillian Mwanri, Karen Hawke, Paul Russell Ward

## Abstract

Access to HIV care service or antiretroviral therapy (ART) is essential for the improvement of health outcomes of people living with HIV (PLHIV) and in reducing HIV transmission and HIV-related deaths. As a part of a qualitative study in Belu, this paper describes the use of traditional treatment and the role of families in determining traditional treatment for their HIV-positive family member as barriers to access to HIV care service or ART among PLHIV. One-on-one in-depth interviews were employed to collect data from 46 PLHIV (26 women and 20 men) and 10 healthcare professionals recruited using the snowball sampling technique. Data analysis was performed using NVivo 12 software and guided by a qualitative data analysis framework. The findings showed that the use of traditional treatment, a well-known cultural practice in Belu, was a barrier to access to HIV care service or ART among PLHIV. The role of family in determining traditional treatment for HIV, supported by the lack of knowledge of ART, effectiveness of traditional medicines in treating other health issues, and social influence of families, neighbours and friends, were also significant barriers to PLHIV’s access to HIV care service or ART. The findings indicate the need for dissemination of HIV care-related information for PLHIV, family and community members to increase their knowledge on the service, ART and its function, and to support and improve access to ART by PLHIV.

## Introduction

Access to HIV care service or antiretroviral therapy (ART) is essential for the improvement of health outcomes of people living with HIV (PLHIV). During the period of 2010 to 2019, ART was reported effective in reducing HIV infections by 23% and AIDS-related deaths by 39% globally [1]. However, during the same period, Indonesia experienced a significant increase of burden of HIV, including an increase of up to 132% infections and 60% AIDS-related deaths, a reflection of the inequitable distribution or limited coverage of ART, late diagnosis, and poor access and adherence to ART [1, 2]. The 2021 national AIDS report shows that of the total number of 427,201 PLHIV in the country, only 63% ever started ART [2]. Of the ones who have started ART, 26.9% have failed to follow up or have stopped the therapy, 18.3% have died, and 53.7% are currently on ART [2].

Globally, previous studies have reported a range of barriers to the access to HIV care services among PLHIV. A limited availability of HIV care services, the shortage of qualified healthcare professionals to deliver the services to PLHIV, long distance travel to healthcare facilities or HIV clinic and the lack of public transportation have been reported as some of the major restrictions for PLHIV to reach and access the services [3-7]. A limited approachability of HIV care services reflected in poor dissemination of information about the services and the poor health literacy of PLHIV about both HIV and HIV care services, have also been reported as factors that influence them to perceive their needs for care [8-10]. Poor health literacy is also reported to influence them in making critical decision about their health, including to seek and access available and appropriate healthcare services [5, 11-13]. The unaffordability of healthcare service-related costs, such as medical and transportation costs, and an inability of PLHIV to afford such costs due to a lack of income, unemployment or poor economic status have also been reported as barriers to their access to the services [5, 9, 14-17]. Other factors such as concerns about confidentiality of HIV status, fear of losing a job if HIV status is known to others, perceived healthy status, lack of time, and psychological burden of undergoing HIV care, are also reported as demotivating factors for their engagement in or access to HIV care services [10, 15, 18-21]. Similarly, perceived, anticipated and external stigma from families, community members and healthcare professionals are also the barriers for PLHIV to engaging in healthcare services, and the supporting factors for the concealment of HIV status and self-isolation of PLHIV [22-27].

Despite a range of barriers to the access to HIV care services as reported in the aforementioned studies, the literature suggests that evidence on the influence of cultural practices (e.g., traditional treatment of HIV using traditional medicines) and social factors (e.g., influence of family members, relatives, friends and neighbours in determining HIV treatment) on the access to HIV care services among PLHIV is still limited. Previous studies in Africa have reported the use of traditional/herbal medicines and its influence on ART adherence among PLHIV [28-32], but none of these studies specifically focused on exploring the use of traditional/herbal medicines as a barrier to the access of HIV care services among PLHIV. For example, studies in Ethiopia and Uganda reported on a common concomitant use of herbal medicines for HIV and ART as a barrier to ART adherence [29-31]. Similarly, a study in Tanzania, Uganda and Zambia reported on consultation with a traditional healer or herbalist as a factor associated with poor ART adherence [28]. Only a recent study in Ethiopia by Gesesew and colleagues, which explored the access of women living with HIV (WLHIV) to HIV care services, reported on the restrictions of traditional healers towards the use of ART as barriers for the women to accessing HIV care services [33]. This paper aims to contribute to the existing knowledge about barriers to access to HIV care services or ART by exploring the use of traditional treatments or medicines for HIV treatment, a well-known cultural practice in Belu, and the role played by the family in determining treatment for their sick family members. In doing this, our findings will be useful information for government and non-governmental institutions and organisations, policy makers and program planners to address the issue through policies and interventions for the better health outcomes of PLHIV in Indonesia and globally.

## Methods

The report of the methods section was guided by consolidated criteria for reporting qualitative studies (COREQ) checklist (S1 Fig) [34]. The checklist contains 32 items that need to be covered to support the explicit and comprehensive reporting of qualitative studies [34].

### Study setting

Belu is a district in East Nusa Tenggara province, located in the eastern part of Indonesia. It shares the border with East Timor with a total population of 204,541 people [35]. The district comprises 12 sub-districts and had three hospitals, 17 community health centers located in each sub-district, 21 sub-community health centers, 23 village health posts, 48 village maternity posts, five private clinics and one HIV clinic where ART is provided. It has a total number of 1,200 PLHIV, but just over half of them (637 people) had accessed HIV care service or ART. Of the ones who had accessed ART, only half regularly accessed and adhered to ART at the time this study was conducted [36]. ART was the only HIV care service available for PLHIV in Belu [36]. The use of traditional treatment for any health issues, including HIV, is also a common practice within families in this area, which might have been a barrier to the access of PLHIV to ART. To the best of our knowledge, there has not been any study exploring the influence of traditional treatment and the social influence of families and friends on access to HIV care service or ART among PLHIV in the context of Belu and Indonesia, and due to feasibility, familiarity and the potential of undertaking the current study successfully, Belu was selected as the study setting.

### Recruitment and data Collection

As part of a larger qualitative study to understand HIV risk factors, impacts and determinants of access to HIV care services among women and men living with HIV in Belu, this paper describes the use of traditional treatment for HIV and the role of family members, friends and neighbours in supporting the use of traditional treatment as barriers to the access of PLHIV to HIV care service or ART. The participants were PLHIV and healthcare professionals (HCPs) recruited using the snowball sampling technique. PLHIV and HCPs were respectively recruited from an HIV clinic and healthcare facilities providing HIV care services in Belu. The study information packs for PLHIV and HCPs were initially distributed to potential participants through an HIV clinic receptionist and healthcare facilities, respectively. Participants who called to confirm their participation were recruited and asked to suggest a preferred time and place for an interview. The initially interviewed participants were also asked to distribute the study information packs to their friends and colleagues who might be willing to participate. This process was iterative, leading to a total of 46 PLHIV (26 women and 20 men) and 10 HCPs participating in this study. Two people withdrew their participation after a few minutes of the start of the interview due to personal reasons and their information was excluded in this report.

One-on-one in-depth interviews were used to collect data from the participants. Interviews with PLHIV and HCPS were respectively conducted in a private room at the HIV clinic and at healthcare facilities where the HCPs worked, which was mutually agreed upon by the field researcher (NKF, male) and each participant. Only the researcher and each participant were present in the interview room. Interviews were conducted in Bahasa for a duration of 35 to 87 minutes and audio recorded. Notes were also taken during the interviews. Pertaining to the topic of traditional treatment, the interviews focused on several key areas, such as PLHIV’s experiences with access to HIV care service or ART, whether or not they regularly access ART, factors that influenced their access to the services or ART, whether or not they used other treatment (e.g., traditional treatment) in addition to ART, the role of their family members in the treatment of HIV, HCPs’ knowledge about barriers to the access to HIV care service or ART by PLHIV and the role of families in HIV treatment for PLHIV. Recruitment of the participants and interviews ceased once information or data provided by the participants in the last few interviews were similar to those of previous participants, an indication of data saturation. Interviews were conducted in Bahasa, the primary language of the participants and the field researcher who can also speak English fluently. The field researcher is a PhD student in public health and had attended training on qualitative research through formal education. The participants (PLHIV) were not offered an opportunity to review their transcript to prevent the possibility of the transcript being received by their family members once it was sent to them in hard copies. This could lead to the breach of the confidentiality of their HIV status, in case they had not disclose it to their family members. No repeated interview was conducted with any participants. No repeated interview was conducted and no established relationship between the researcher and any participants prior to the interviews.

### Data analysis

The audio recordings were transcribed verbatim and manually using a laptop by the first author (NKF). This was started alongside the data collection process. Emotions and notes made during the interview with each participant were integrated into each individual transcript at the transcription stage. Data analysis was performed using NVivo 12 software and guided by the five steps introduced in qualitative data analysis framework by Ritchie and Spencer [37], including: (i) familiarisation with the data or transcripts, (ii) identification of a thematic framework, (iii) indexing the data, (iv) charting data, and (v) mapping and interpretation of the data. In the first step, each transcript was read line by line repeatedly for familiarisation purposes, and ideas related to the barriers facing PLHIV in their access to HIV care services or ART were highlighted. Comments and labels were made to data extracts of each individual transcript to search for meanings, patterns and ideas. In the second step, a thematic framework was identified. This was performed by writing down key issues and concepts that had been highlighted in the first step. These key issues and concepts were then used to form the thematic framework. This was an iterative process and involved changing and refining the themes. In the third step, data were indexed or coded which was started by creating open coding to each individual transcript, where data extracts of each transcript were given a code or node. This process led to a collection of a long list of open codes or nodes from each individual transcript. This was followed by close coding where similar or redundant codes or nodes were identified and grouped together to reduce the length of the coding list, and then codes or nodes that appeared to form the same themes or sub-themes were grouped together. In the fourth step, the data were charted by arranging a thematic framework in a summary of chart. This was performed by reorganising codes or nodes that had been created in each individual transcript and putting them together under each theme in a chart. This also enable the comparison across the transcripts or within each individual transcript. Finally, the data were mapped and interpreted as a whole [37, 38].

### Ethical consideration

The study was approved by Social and Behavioural Research Ethics Committee, Flinders University, Australia (No. 8286), and Health Research Ethics Committee, Duta Wacana Christian University, Indonesia (No. 1005/C.16/FK/2019). All the study participants were informed about these ethics approvals prior to commencing the interviews. They were informed about the purpose of the study through the study information packs and by the research prior to the interview. They were also advised that their participation was voluntary, and that they had the rights to withdraw their participation for any reasons before or during the interview without any consequences. They were assured that information they provided during the interview will be treated confidentially and anonymously by assigning each participant a specific letter and number. This helps to prevent the possibility of linking back the data to any individual in the future. Before commencing the interviews, each participant was provided with an informed consent to sign and return to the research.

## Results

### Traditional treatment of HIV

#### Traditional treatment as a cultural practice

The use of traditional medicines to treat any kinds of diseases, including HIV/AIDS was common among community members in Belu. Both PLHIV and HCPs acknowledged that the use of traditional medicines provided by traditional healers had been a well-known cultural practice within communities, passed down from one generation to another:

> *“It has been our culture since long time ago, from our ancestors that if people get sick then they would firstly seek for traditional medicines” (FP15)*.
>
> *“The use of traditional medicines for HIV treatment is very common among PLHIV in Belu. I am sure that most PLHIV who have known their HIV status but do not start antiretroviral therapy are taking traditional medicines” (HCP1)*.

The cultural practice of traditional treatment was reported to influence or delay the acceptability of and access to HIV care services among both women and men living with HIV in Belu. It influenced the initiation of and retention in HIV treatment or ART. Nearly half of the women and men interviewed described that they did not access the HIV care service or ART straightaway following their HIV diagnosis or stopped the ART due to undergoing traditional treatment using traditional medicines. The same information was also provided by all HCPs:

> *“After the HIV diagnosis, the doctor told us (the woman and her spouse) that the medicines for HIV treatment are available at this hospital (HIV clinic) but we did not access the medicines directly. We did the treatment using traditional medicines provided by a traditional healer in XX (name of a place)” (FP8)*.
>
> *“After the diagnosis I accessed the therapy (ART) and then once I finished the medicines (first month), I switched to traditional medicines. I came back here (to restart ART) because my physical condition was getting weaker” (MP9)*.
>
> *“There are patients who do not use traditional medicines straightaway after the HIV diagnosis even though we (healthcare professionals) have talked to them and encouraged them to start antiretroviral therapy. There are also patients who have started ARV therapy for a few months and then quitted and switched to traditional medicines” (HCP4)*.

#### Health and economic consequences of traditional treatment of HIV

The use traditional medicines for HIV treatment caused negative impacts on the health of PLHIV. Both women and men living with HIV, who previously underwent traditional treatment, described that traditional treatment worsened their health, which was a supporting reason for them to start or restart ART. Similarly, the HCPs also commented undergoing traditional treatment of HIV or switching from ART to traditional treatment often deteriorated physical health condition of PLHIV:

> *“My spouse and I took a traditional medicine, but my spouse’s condition got worse, so we went back to hospital (to do continue treatment with ART, her spouse died from AIDS)” (FP4)*.
>
> *“There were also patients who have started the therapy (ART) for a few months and then quitted and switched to traditional medicines. Some of these patients came back to this clinic to restart the therapy once their physical and health conditions gradually got worse” (HCP3)*.

The use of traditional medicines was reported to have financial consequences for PLHIV in Belu as reported by both PLHIV and HCPs in this study. Some participants, both women and men living with HIV, acknowledged that they had to pay a certain amount of money and give sacrificial animal to traditional healers who provided the traditional medicines. The amount of money and the animal seemed to differ from one traditional healer to another and had to be provided prior to the commencement of the treatment. Meanwhile some other participants revealed that they had the traditional treatment for free as the traditional healers were in their families:

> *“The use of traditional medicines for treatment of any kinds of disease is very common here, but it is costly. Patients have to pay some amount of money and take with them a chicken or pig or goat as sacrificial animal. I heard that to get traditional medicines from Marry (pseudonym of the traditional healer), a patient has to pay five million rupiahs and give a chicken or pig to her” (MP16)*.
>
> *“My spouse and I took traditional medicines from three different traditional healers and wasted a lot of money which was up to two million rupiahs for each of them and one goat, but we did not get better” (FP5)*.
>
> *“I used traditional medicines from my family members, they knew that I am infected with HIV, and offered the medicines to me. I did not pay them, they are my family, they just wanted to help me and did not ask for money” (FP7)*.

The desire to recover faster and completely cured from HIV, and distrust in HIV test results, were reported as barriers to the acceptance of medical treatment or ART among PLHIV.

These were acknowledged by the participants from both groups as supporting factors for the decision of PLHIV and their families not to access HIV care service or ART or to switch from ART to traditional treatment:

> *“To get the traditional medicines and undergo the treatment with the healer, we (the man and his spouse who is also HIV-positive) had to pay a lot of money to him (the healer) and give him a goat. It was very expensive, but we had to do it because we wanted to get better. We spent about fifteen million rupiahs for this treatment. This amount had to be paid at once before the start of the treatment, only once. My family members helped me with the payment” (MP11)*.
>
> *“One of the reasons they or their families decide to use traditional medicines or switch from antiretroviral therapy to traditional treatment of HIV is because of the desire to get better faster or full recovery. Some patients told me that they wanted to get cured and strong faster, that is why they switched to traditional medicines, even though we have told them that they will not be completely cured of HIV and therefore they have to take antiretroviral medicines every day for the rest of their lives” (HCP9)*.

### The role of family in the use of traditional medicines for HIV treatment

#### Family decisions for traditional treatment of HIV

The participants, both PLHIV and HCPs, reported that family members had a crucial role in determining the treatment for PLHIV. Several female and male participants (PLHIV) commented that they underwent HIV treatment using traditional medicines due to being asked by their family members. Such an influence of family members was also reported by HCPs:

> *“I had started taking the ART, but my family members asked me to take a traditional medicine. So, I just did what they asked me to do because they said that the traditional medicine is good to treat HIV” (MP10)*.
>
> *“After he (her spouse) was diagnosed with HIV, at first the doctor gave him cotrimoxazole for two weeks, but they (her spouse’s family) told him to take traditional medicines. …. They asked him and me to use the traditional medicines for bathing as well. My spouse and I took the traditional medicines, but after a while my spouse’s condition got worse, so we went back to hospital (to start ART but her spouse died)” (FP4)*.
>
> *“I see that family members have a dominant role in determining treatment for the sick ones (HIV-positive). Often people are diagnosed with HIV while they are in a severe condition and being admitted to hospital. So, once they left the hospital, many of them switched to traditional medicines because their family members asked them to do so and then stopped taking antiretroviral medicines. Family members are the ones who look for traditional medicines for them (PLHIV)” (HCP10)*.

Both PLHIV and HCPs described that family members’ decisions to use traditional medicines for HIV treatment of their sick family member was influenced by their experience of the effectiveness of traditional medicines in treating other health issues. Several PLHIV described that their family members were regular, long-term users of traditional medicines, and therefore asked them to use traditional medicines for HIV treatment. This also seemed to be supported by the lack of knowledge among their family members about ART provided in the HIV clinic:

> *“My family members are very familiar with traditional medicines and these are number one medicines for them. Every time they feel sick or unwell, they use traditional medicines to treat their body. So, they recommended me to use traditional medicines to treat HIV as well. They do not know anything about medical treatment like these (showing her ARV medicines she just collected)” (FP5)*.
>
> *“In general, (HIV) patients come from families with low level education. Their family members do not have knowledge about ART, do not understand about the function of the therapy to suppress viral load. Besides, I believe their family members have seen and experienced healing from certain ailments or health issues due to the use of traditional medicines. I think, these are also the reasons why many families rely on traditional medicines for the treatment of their HIV-positive family member” (HCP3)*.

Besides, the stories of some HCPs and PLHIV interviewed in this study showed that being physically weak and taken care of by their family members, and a lack of knowledge about ART, appeared to be personal-related supporting factors that made them accept and follow the recommendations of their family members for the use of traditional medicines:

> *“Once we (the woman and her spouse) were tested positive with HIV, our physical conditions were weak already, our families took care of us. My family members asked us to use traditional medicines. So, we just used them, my family members sent them to us every month. We just listened to what they said and used the traditional medicines because we wanted this HIV to go away” (FP23)*.
>
> *“It is true that family members play a very important role in HIV treatment for HIV patients. If their family members provide traditional medicines, then they (PLHIV) will definitely take the medicines or switch to the traditional treatment. There is no way for them to refuse because they are taken care of by family members, and they are sick” (HCP7)*.

#### Social influence on the use of traditional treatment of HIV

Extended family members and neighbours were also reported to have an influence on the decision of the participants’ family members for the use of traditional medicines to treat HIV infection. Providing information about traditional medicines for HIV treatment, and encouraging the participants’ family members about the effectiveness of traditional medicines to treat HIV infection, were in some instances reflecting the influences of others towards the participants’ family members regarding the use of traditional medicines:

> *“My family members encouraged us (the women and her spouse who died from AIDS) to use traditional medicines because they were encouraged by our extended family members and neighbours that traditional medicines can cure HIV (FP8)*.
>
> *“We (the man and his spouse) know about a traditional medicine for HIV from a friend of mine. He came to my house and talked to my spouse and me about it. My spouse was convinced with his story that the traditional medicine is very good and has cured people from this disease (HIV infection). Then my spouse encouraged me to switch (from ART) to that traditional medicine” (MP9)*.

## Discussion

Increased HIV transmissions and AIDS-related deaths in many countries and regions globally have been associated with poor access to HIV care services and poor adherence to ART (UNAIDS, 2020). This paper describes the influence of the use of traditional treatment of HIV and the role of families, friends and neighbours in determining HIV treatment among PLHIV, which have not been addressed in previous literature on barriers to ART adherence [39-42].

The current study suggests that participants’ (PLHIV) access to HIV care service or ART was influenced by the use of traditional treatment of HIV provided by traditional healers, a well-known cultural practice which has been passed down from one generation to another within communities in Belu. Such influence was reflected in the fact that some ‘decided’ not to access HIV care services or to switch to traditional treatment after ART initiation or to treat HIV using traditional medicines at the first place. Availability and approachability of traditional medicines for any health issues were indicated as the supporting factors for the use of traditional treatment for HIV, which was a barrier to the access to HIV care service or ART among the participants (PLHIV). These support the constructs of access to healthcare service framework [9], suggesting availability of a healthcare service or treatment and approachability or how well-known the information about the service or treatment is among people in health needs as some of the dimensions determining or influencing accessibility of the service. The current findings extend the understanding of the use of traditional medicines or therapies and the restrictions traditional healers towards ART, which have been reported in previous studies in Africa as barriers to ART adherence [28-32]. The current study also suggests that the use of traditional medicines for HIV treatment had a negative financial impact on some participants as the service costed large amounts of money and required sacrificing animals.

Evidence from the current study suggests that the role of family members in determining HIV treatment using traditional medicines from traditional healers was also a significant barrier to the access to and initiation of ART among the participants (PLHIV) in Belu. Some PLHIV and healthcare providers reported that the use of traditional medicines by PLHIV for HIV treatment was mainly the decision made by family members, such as parents, siblings and spouses, which hindered or led to the delay of access to HIV care services, ART initiation and non-adherence to ART. The regular use of traditional medicines within families and the positive previous experience of the effectiveness of traditional medicines in treating other health issues, were the underlying reasons for family decisions to use of traditional treatment over ART for their infected family member. Similarly, the findings suggest that a lack of understanding among both participants (PLHIV) and their family members about the function of ART to supress the viral load, led to the ‘choice’ of traditional medicines over antiretroviral medicines for HIV treatment in the early stage of the participants’ HIV diagnosis. This was also reflected in the fact that access to HIV care services or ART was a last choice for participants (PLHIV), and this was made once traditional treatment was felt to be ineffective and their physical and health conditions were getting worse. These are not dissimilar to previous findings which reported insufficient or incorrect knowledge about ART as a barrier to the access and adherence to ART among PLHIV [41], and positive relationships between a lack of ART knowledge and less family support for ART access and adherence [43]. In other words, the current study indicates a lack of family support for PLHIV prior to or in the early stages of ART as a barrier to their access to HIV care services and ART initiation or adherence [39-42].

The current findings also suggest that the social influence of extended families, neighbours and friends on family decisions for the use of traditional medicines from traditional healers for HIV treatment was another barrier for access to HIV care services and ART initiation among PLHIV in Belu. This was reflected in the comments of some PLHIV in Belu that explained about the encouragement, provision of traditional medicines or information about traditional treatment by extended families, neighbours and friends as factors that influenced their family’s decisions for the use of traditional medicines from traditional healers for HIV treatment. The findings add further evidence to a previous study’s findings [43] which have reported that social factors, such as gender norms, social status and stigma affected family support for PLHIV and their access to ART. In addition, evidence from the current study also demonstrates that the poor physical and health conditions of PLHIV, and their dependency on family support for their daily needs and healthcare influenced their ability to make decisions about their own health treatment, hence indulging family decisions for traditional treatment, which negatively affected their access to ART.

### Study limitations and strengths

PLHIV who participated in this study were recruited from an HIV clinic Belu, whilst some of them may have stopped ART at some point, they are still engaged in care. We did not include PLHIV who are disengaged from care, who may have had different stories to tell about the impact of family or traditional medicines on their use of ART. However, to our knowledge, this is the first qualitative inquiry exploring the influence of traditional treatment and the role of families on the access to HIV care services among PLHIV in the context of Indonesia, hence the findings are useful information for the development of interventions that address barriers to access to HIV care services facing PLHIV.

## Conclusions

The paper reports the use of traditional treatments and the role of family members in determining treatment for their HIV-positive family member as barriers to the access to HIV care services among PLHIV in Belu. It shows that the use of traditional medicines in treating any kind of health issues, including HIV is a well-known cultural practice within communities in Belu, which influenced participants’ (PLHIV) access to HIV care services and ART adherence. It also reports a strong family role in determining the use of traditional treatment for an HIV-positive family member, which was supported by a lack of knowledge about ART, positive experience of the effectiveness of traditional medicines in treating other health issues and the social influence of other. The findings indicate the need for dissemination of ART-related information for PLHIV, family and community members to increase their knowledge on ART and its function, support and improve the access to HIV care services among PLHIV. Future studies exploring cultural and family-related barriers to the access to HIV care services among PLHIV in other settings are recommended.

## Data Availability

All relevant data are within the manuscript.

## Acknowledgement

Not applicable.

---

S1 Fig.: COREQ checklist.

## Reference

1. UNAIDS. UNAIDS data Geneva, Switzerland: Joint United Nations Programme on HIV/AIDS; 2020.

2. Kementerian Kesehatan RI. Laporan Situasi Perkembangan HIV/AIDS dan PIMS di Indonesia, Triwulan IV Tahun 2020. Jakarta, Indonesia: Kementerina Kesehatan Republik Indonesia. Available at: https://siha.kemkes.go.id/portal/files_upload/Laporan_TW_IV_2020.pdf; 2021.

3. Arcaya MC, Arcaya AL, Subramanian SV. Inequalities in health: definitions, concepts, and theories. Global Health Action. 2015;8:1–12.

4. Deaton A. Health, Inequality, and Economic Development. Journal of Economic Literature. 2003;XLI:113–58.

5. Fauk NK, Merry MS, Putra S, Sigilipoe MA, Crutzen R, Mwanri L. Perceptions among transgender women of factors associated with the access to HIV/AIDS-related health services in Yogyakarta, Indonesia. PLoS ONE. 2019;14(8):1–17.

6. Fauk NK, Sukmawati AS, Wardojo SSI, Teli M, Bere YK, Mwanri L. The Intention of Men Who Have Sex With Men to Participate in Voluntary Counseling and HIV Testing and Access Free Condoms in Indonesia. American Journal of Men’s Health. 2018;Special Section:1–10.

7. Whitehead M. The concepts and principles of equity and health. International Journal of Health Services. 1992;22:429–45.

8. Keleher H, Hagger V. Health Literacy in Primary Health Care. Australian Journal of Primary Health. 2007;13(2):24–30.

9. Levesque J-F, Harris MF, Russell G. Patient-centred access to health care: conceptualising access at the interface of health systems and populations. Int J Equity Health. 2013;12(18):1–9.

10. Li H, Wei C, Tucker J, Kang D, Liao M, Holroyd E, et al. Barriers and facilitators of linkage to HIV care among HIV-infected young Chinese men who have sex with men: a qualitative study. BMC Health Services Research. 2017;17(214):1–8.

11. Choi KH, Lui H, Guo Y, Han L, Mandel JS. Lack of HIV testing and awareness of HIV infection among men who have sex with men, Beijing, China. AIDS Education and Prevention. 2006;18:33–43.

12. Fauk NK, Kustanti CY, Liana DS, Indriyawati N, Crutzen R, Mwanri L. Perceptions of Determinants of Condom Use Behaviors Among Male Clients of Female Sex Workers in Indonesia: A Qualitative Inquiry. Am J Mens Health. 2018;Special Section:1–10.

13. Mohlabane N, Tutshana B, Peltzer K, Mwisongo A. Barriers and facilitators associated with HIV testing uptake in South African health facilities offering HIV Counselling and Testing. Health SA Gesondheid. 2016;21:86–95.

14. Shengelia B, Murray CJL, Adams OB. Beyond access and utilization: defining and measuring health system coverage. In: Murray CJL, Evans DB, editors. Health SystemsPerformance Assessment Debates, methods and empiricism. Geneva: World Health Organization; 2003. p. 221–34.

15. Nakanwagi S, Matovu JB, Kintu BN, Kaharuza F, Wanyenze RK. Facilitators and Barriers to Linkage to HIV Care among Female Sex Workers Receiving HIV Testing Services at a Community-Based Organization in Periurban Uganda: A Qualitative Study. Journal of Sexually Transmitted Diseases. 2016;2016:1–9.

16. Salkever DS. Accessibility and the demand for preventive care. Social Science and Medicine. 1976;10:469–75.

17. Local Burden of Disease HIV Collaborators. Mapping subnational HIV mortality in six Latin American countries with incomplete vital registration systems. BMC Medicine. 2021;19(1):4.

18. Liu Y, Osborn CY, Qian H-Z, Yin L, Xiao D, Ruan Y, et al. Barriers and Facilitators of Linkage to and Engagement in HIV Care Among HIV-Positive Men Who Have Sex with Men in China: A Qualitative Study. AIDS Patient Care and STDs. 2016;30(2):70–7.

19. Nyato D, Nnko S, Komba A, Kuringe E, Plotkin M, Mbita G, et al. Facilitators and barriers to linkage to HIV care and treatment among female sex workers in a community-based HIV prevention intervention in Tanzania: A qualitative study. PLoS ONE. 2019;14(11):1–14.

20. Tso LS, Best J, Beanland R, Doherty M, Lackey M, Ma Q, et al. Facilitators and Barriers in HIV Linkage to Care Interventions: A Qualitative Evidence Review. AIDS. 2016;30(10):1639–53.

21. Sartorius B, VanderHeide JD, Yang M, Goosmann EA, Hon J, Haeuser E, et al. Subnational mapping of HIV incidence and mortality among individuals aged 15–49 years in sub-Saharan Africa, 2000–18: a modelling study. The Lancet HIV. 2021;8(6):e363–e75.

22. Geter A, Herron AR, Sutton MY. HIV-Related Stigma by Healthcare Providers in the United States: A Systematic Review. AIDS Patient Care and STDs. 2018;32(10):418–24.

23. Mahamboro DB, Fauk NK, Ward PR, Merry MS, Siri TA, Mwanri L. HIV Stigma and Moral Judgement: Qualitative Exploration of the Experiences of HIV Stigma and Discrimination among Married Men Living with HIV in Yogyakarta. International Journal of Environmental Research and Public Health. 2020;17(363):1–15.

24. Stangl AL, Earnshaw VA, Logie CH, van Brakel W, Simbayi LC, Barré I, et al. The Health Stigma and Discrimination Framework: a global, crosscutting framework to inform research, intervention development, and policy on health-related stigmas. BMC Medicine. 2019;17(31):1–13.

25. Vorasane S, Jimba M, Kikuchi K, Yasuoka J, Nanishi K, Durham J, et al. An investigation of stigmatizing attitudes towards people living with HIV/AIDS by doctors and nurses in Vientiane, Lao PDR. BMC Health Services Research. 2017;17(125):1–13.

26. Fauk NK, Hawke K, Mwanri L, Ward PR. Stigma and Discrimination towards People Living with HIV in the Context of Families, Communities, and Healthcare Settings: A Qualitative Study in Indonesia. International Journal of Environmental Research and Public Health. 2021;18(5424):1–17.

27. Fauk NK, Ward PR, Hawke K, Mwanr L. HIV Stigma and Discrimination: Perspectives and Personal Experiences of Healthcare Providers in Yogyakarta and Belu, Indonesia. Frontiers in Medicine. 2021;8:625.

28. Denison JA, Koolec O, Tsui S, Menten J, Torpey K, van Praag E, et al. Incomplete adherence among treatment-experienced adults on antiretroviral therapy in Tanzania, Uganda and Zambia. AIDS. 2015;29(3):361–71.

29. Gurmu AE, Teni FS, Tadesse WT. Pattern of Traditional Medicine Utilization among HIV/AIDS Patients on Antiretroviral Therapy at a University Hospital in Northwestern Ethiopia: A Cross-Sectional Study. Evidence-Based Complementary and Alternative Medicine. 2017;2017:1–7.

30. Haile KT, Ayele AA, Mekuria AB, Demeke CA, Gebresillassie BM, Erkub DA. Traditional herbal medicine use among people living with HIV/AIDS in Gondar, Ethiopia: Do their health care providers know? Complementary Therapies in Medicine. 2017;35:14–9.

31. Lubinga SJ, Kintu A, Atuhaire J, Asiimwe S. Concomitant herbal medicine and Antiretroviral Therapy (ART) use among HIV patients in Western Uganda: A crosssectional analysis of magnitude and patterns of use, associated factors and impact on ART adherence. AIDS Care. 2012;24(11):1375–83.

32. Peltzer K, Preez NF-D, Ramlagan S, Fomundam H, Anderson J, Chanetsa L. Antiretrovirals and the use of traditional, complementary and alternative medicine by HIV patients in Kwaszulu-Natal, South Africa: a longitudinal study. African Journal of Traditional, Complementary and Alternative Medicine. 2011;8(4):337–45.

33. Gesesew H, Lyon P, Ward P, Woldemichael K, Mwanri L. “Our Tradition Our Enemy”: A Qualitative Study of Barriers to Women’s HIV Care in Jimma, Southwest Ethiopia. International Journal of Environmental Research and Public Health. 2020;17(833).

34. Tong A, Sainbury P, Craig J. Consolidated criteria for reporting qualitative research (COREQ): a 32-item checklist for interviews and focus groups. International Journal for Quality in Health Care. 2007;19(6):349–57.

35. BPS Kabupaten Belu. Kabupaten Belu Dalam Angka (Belu Regency in Figures). Atambua, Indonesia: Badan Pusat Statistik Kabupaten Belu; 2021.

36. Dinkes Belu. Laporan Perkembangan Kasus HIV/AIDS di Belu. Atambua: Dinas Kesehatan Kabupaten Belu; 2020.

37. Ritchie J, Spencer L. Qualitative Data Analysis for Applied Policy Research. In: Bryman A, Burgess RG, editors. London: Routledge; 1994. p. 173–94.

38. Fauk NK, Ward PR, Hawke K, Mwanri L. Cultural and religious determinants of HIV transmission: a qualitative study with people living with HIV in Belu and Yogyakarta, Indonesia. PLoS One. 2021;16(11):e0257906.

39. Bazzi AR, Drainoni M-L, Biancarelli DL, Hartman JJ, Mimiaga MJ, Mayer KH, et al. Systematic review of HIV treatment adherence research among people who inject drugs in the United States and Canada: evidence to inform pre-exposure prophylaxis (PrEP) adherence interventions. BMC Public Health. 2019;19(31):1–10.

40. Croomea N, Ahluwalia M, Hughes LD, Abasa M. Patient-reported barriers and facilitators to antiretroviral adherence in sub-Saharan Africa. AIDS. 2017;31:995–1007.

41. Engler K, Lènàrt A, Lessard D, Toupin I, Lebouché B. Barriers to antiretroviral therapy adherence in developed countries: a qualitative synthesis to develop a conceptual framework for a new patient-reported outcome measure. AIDS Care. 2018;30(Sup1):17–28.

42. Shubber Z, Mills EJ, Nachega JB, Vreeman R, Freitas M, Bock P. Patient-Reported Barriers to Adherence to Antiretroviral Therapy: A Systematic Review and Meta-Analysis. PLoS Med. 2016;13(11):e1002183.

43. Campbell L, Masquillier C, Thunnissen E, Ariyo E, Tabana H, Sematlane N, et al. Social and Structural Determinants of Household Support for ART Adherence in Low- and Middle-Income Countries: A Systematic Review. Int J Environ Res Public Health. 2020;17(3808):1–28.

